# The Impact of Short Sleep Duration on Depression, Mental Health Days, and Physical Health

**DOI:** 10.1101/2025.03.06.25323545

**Authors:** Mojisola Fasokun, Oluwasegun Akinyemi, Fadeke Ogunyankin, Phiwinhlanhla Ndebele-Ngwenya, Kaelyn Gordon, Seun Ikugbayigbe, Uzoamaka Nwosu, Mariam Michael, Kakra Hughes, Temitope Ogundare

## Abstract

**Introduction:** Sleep is essential for mental and physical well-being, yet a significant proportion of U.S. adults experience insufficient sleep (<7 hours per night). Short sleep duration has been associated with an increased risk of mental health disorders and poor physical health, but limited studies have quantified these associations.

**Objective:** This study examines the impact of short sleep duration on depression, self-reported poor mental health days, and poor physical health days.

**Methodology:** Data were obtained from the Behavioral Risk Factor Surveillance System (BRFSS) (2016–2023). Sleep duration was categorized as short sleep (<7 hours, coded as 1) or adequate sleep (≥7 hours, coded as 0). The primary outcomes were depression diagnosis, poor mental health days, and poor physical health days. Inverse Probability Weighting (IPW) was used to estimate the Average Treatment Effect (ATE), adjusting for demographic and socioeconomic factors.

**Results:** Short sleep duration was associated with a 5.6% increased risk of depression (ATE = 0.056, p < 0.001), 2.24 additional poor mental health days per month (ATE = 2.24, p < 0.001), and 1.8 more poor physical health days per month (ATE = 1.76, p < 0.001).

**Conclusion:** Short sleep duration significantly increases the risk of depression and worsens mental and physical health. Public health interventions promoting sleep hygiene are needed to mitigate these effects and improve overall well-being.

## Introduction

Sleep is a fundamental biological necessity essential for overall health, cognitive function, and emotional well-being(1, 2). Despite its critical role, sleep deprivation is a growing public health concern in the United States(3, 4). The Centers for Disease Control and Prevention (CDC) recognizes insufficient sleep as a public health epidemic, with a significant proportion of adults failing to meet the recommended minimum of seven hours of sleep per night(5). National surveys indicate that approximately one in three adults in the U.S. experience short sleep duration (<7 hours per night), with even higher prevalence among certain sociodemographic groups, including racial minorities, low-income individuals, and those with demanding work schedules(6–8). The consequences of sleep deprivation extend far beyond individual health, significantly impacting work productivity, increasing absenteeism, and contributing to workplace accidents(9, 10). According to estimates, sleep deprivation costs the U.S. economy billions of dollars annually in lost productivity and healthcare expenditures(11). The economic burden extends to American taxpayers, as insufficient sleep contributes to higher medical costs associated with sleep-related disorders, increased emergency department visits, and treatment of mental health conditions(12). Furthermore, sleep deprivation is closely linked to diminished quality of life, increased risk of chronic diseases, and impaired daily functioning, ultimately affecting the overall well-being of individuals and society at large(13, 14).

Multiple factors contribute to short sleep duration in the U.S., including work-related stress, shift work, socioeconomic disparities, lifestyle habits, and mental health disorders(15–17). Modern lifestyle changes, including excessive screen time, irregular sleep schedules, and high levels of occupational stress, further exacerbate the prevalence of insufficient sleep(18). In addition, underlying health conditions such as anxiety, depression, and chronic pain contribute to sleep disturbances, creating a vicious cycle where inadequate sleep exacerbates these conditions.

Insufficient sleep has profound effects on both mental and physical health, increasing susceptibility to mood disorders such as depression and anxiety while also reducing motivation and capacity for physical activity(19, 20). Individuals experiencing short sleep duration are more likely to report poor mental health days, decreased emotional resilience, and higher levels of stress. In addition, reduced sleep duration is associated with lower levels of physical activity, increasing the risk of obesity, cardiovascular diseases, and metabolic disorders(21–23).

Understanding the relationship between insufficient sleep and these health outcomes is essential for informing public health interventions and policy decisions aimed at improving overall well-being.

While existing literature has established associations between sleep deprivation and health outcomes(24, 25), there is a critical gap in quantifying the direct impact of short sleep duration on mental and physical health, particularly in terms of its effects on depression, poor mental health days, and physical activity. Previous studies have primarily focused on specific populations or relied on self-reported sleep duration without adequately addressing the broader public health implications(26, 27). This study aims to fill this gap by providing a comprehensive analysis of how short sleep duration influences mental and physical health outcomes.

The primary objective of this study is to quantify the impact of insufficient sleep (<7 hours per night) on depression, self-reported poor mental health days, and physical activity levels. By examining these relationships, we seek to provide data-driven insights that can inform targeted interventions aimed at improving sleep hygiene and mitigating the adverse effects of sleep deprivation on population health.

## Methodology

### Study Design and Data Source

This study utilizes data from the Behavioral Risk Factor Surveillance System (BRFSS)(28, 29), a nationally representative, cross-sectional survey conducted annually by the Centers for Disease Control and Prevention (CDC). The BRFSS collects health-related data from U.S. adults through telephone-based interviews, providing extensive information on health behaviors, chronic conditions, and healthcare access(30). Data from 2016 to 2023 were used for this study, ensuring a broad temporal scope to assess trends and associations between sleep duration, mental health outcomes, and physical activity levels.

### Primary Explanatory Variable: Sleep Duration

The primary explanatory variable in this study is self-reported sleep duration, measured in hours per night. Sleep duration was dichotomized into a binary variable, where individuals who reported sleeping less than seven hours per night were categorized as experiencing short sleep duration (coded as 1), while those who reported sleeping seven or more hours per night were categorized as having adequate sleep duration (coded as 0). This classification aligns with recommendations from the American Academy of Sleep Medicine (AASM) and CDC guidelines, which define fewer than seven hours of sleep as insufficient for optimal health.

### Outcome Variables

The primary outcome variables include depression diagnosis, poor mental health days, and physical activity levels. Depression diagnosis was determined using self-reported responses to the BRFSS question, “(Ever told) (you had) a depressive disorder (including depression, major depression, dysthymia, or minor depression)?” Responses were categorized as either yes (coded as 1) or no (coded as 0). Poor mental health days were assessed using the BRFSS item, “For how many days during the past 30 days was your mental health not good?” Responses ranged from 0 to 30 days, representing the self-reported number of days with poor mental health. Physical activity levels were based on BRFSS responses regarding engagement in moderate or vigorous physical activity in the past month. Individuals who reported engaging in regular physical activity were coded as 1 (physically active), while those who did not were coded as 0 (physically inactive).

### Covariates and Confounder Adjustment

To control for potential confounding factors, the analysis adjusted for key demographic and socioeconomic characteristics, including age, sex, race/ethnicity, marital status, employment status, education level, metropolitan versus non-metropolitan residence, state of residence, and survey year. These covariates were included to ensure a more accurate estimation of the association between sleep duration and health outcomes by accounting for variations in lifestyle, socioeconomic status, and access to healthcare services.

### Statistical Analysis

For statistical analysis, the study employed Inverse Probability Weighting (IPW) to estimate the Average Treatment Effect (ATE) of short sleep duration on depression, poor mental health days, and physical activity levels. The IPW approach accounted for potential confounding by balancing covariates between individuals with short and adequate sleep durations. A logistic regression model was used for binary outcomes such as depression diagnosis and physical activity status, while a negative binomial regression model was applied to analyze poor mental health days, given that this outcome represents count data with overdispersion. To ensure national representativeness, BRFSS-provided survey weights were applied, and robust standard errors were clustered at the state level to account for intra-state correlation. In addition, state and year fixed effects were included in the model to adjust for regional and temporal differences in sleep duration and health outcomes.

### Ethical Considerations

This study utilized publicly available, de-identified data from the BRFSS, which is collected and managed by the Centers for Disease Control and Prevention (CDC). As a secondary analysis of de-identified survey responses, this study did not involve human participants directly and did not require Institutional Review Board (IRB) approval. According to CDC guidelines, BRFSS data are fully anonymized before being made publicly available, and the CDC obtains informed consent from participants at the time of data collection.

Since the dataset was fully de-identified and publicly accessible, participant consent for this secondary analysis was not required, and the need for consent was waived by the CDC. This study adheres to ethical research standards and complies with all relevant data use policies.

## RESULTS

Table 1 presents the baseline characteristics of the study population before and after propensity score matching. The unmatched sample included 3,428,251 respondents, of which 565,626 (16.5%) reported sleeping fewer than seven hours per night, while 2,862,625 (83.5%) reported sleeping at least seven hours per night. After matching, the final analytic sample consisted of 211,339 individuals in each group, achieving a well-balanced cohort for comparative analysis.

**Table 1:**
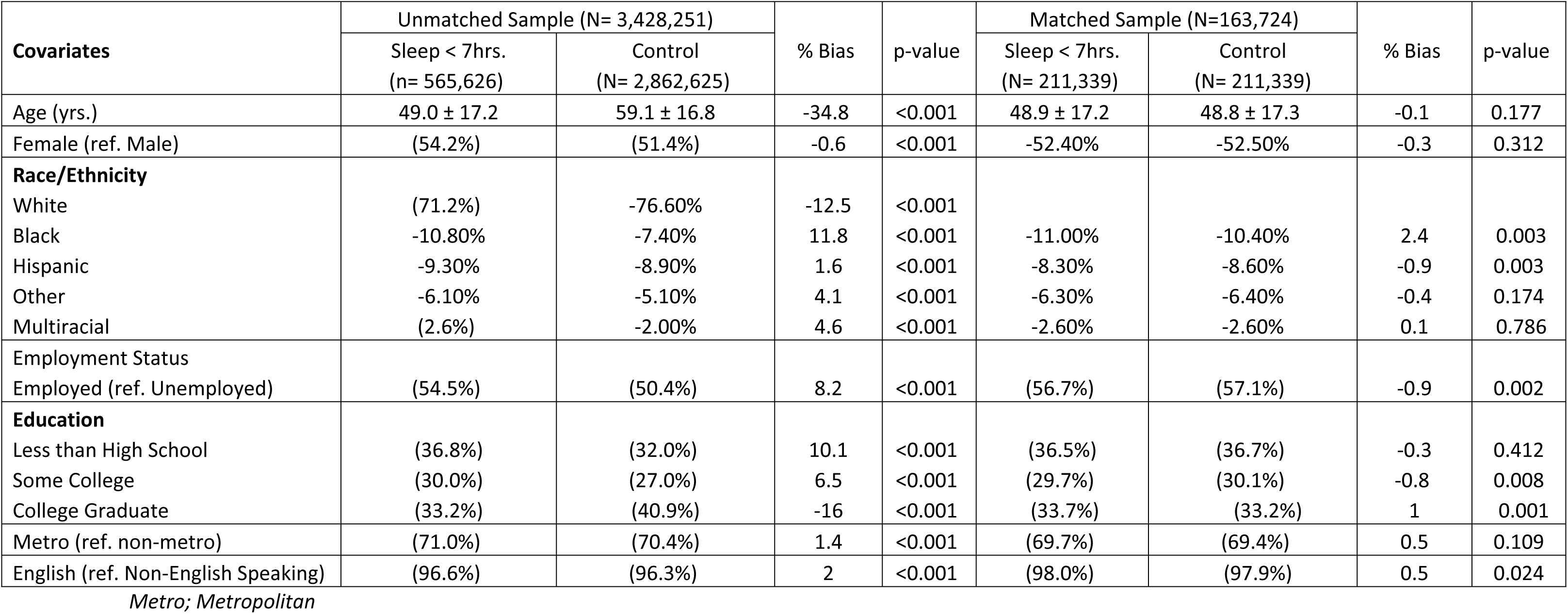
Baseline Covariate Balance for Unmatched and Matched Samples.

In the unmatched sample, individuals who reported short sleep duration (<7 hours) were younger on average (49.0 ± 17.2 years) compared to those with adequate sleep (59.1 ± 16.8 years), with a significant bias of −34.8% (p < 0.001). After matching, the mean ages were balanced between groups (48.9 ± 17.2 vs. 48.8 ± 17.3 years), reducing the bias to −0.1% (p = 0.177).

Sex distribution also varied in the unmatched cohort, with a slightly higher proportion of females among those with short sleep duration (54.2%) compared to controls (51.4%), with a bias of −0.6% (p < 0.001). This difference was substantially reduced after matching (52.4% vs. 52.5%, bias −0.3%, p = 0.312).

Race and ethnicity were significantly different between groups in the unmatched sample. The proportion of White individuals was lower in the short sleep group (71.2%) compared to the control group (76.6%), with a bias of −12.5% (p < 0.001). Black individuals were more prevalent in the short sleep group (10.8% vs. 7.4%, bias 11.8%, p < 0.001), while smaller differences were observed for Hispanic (9.3% vs. 8.9%) and other racial groups (6.1% vs. 5.1%). After matching, race distributions were well-balanced across groups, with minimal residual biases.

Regarding employment status, individuals with short sleep duration were more likely to be employed (54.5%) compared to the control group (50.4%), with a bias of 8.2% (p < 0.001). However, after matching, employment rates were nearly identical between groups (56.7% vs. 57.1%, bias −0.9%, p = 0.002).

Differences in educational attainment were also observed in the unmatched sample. A higher proportion of those with short sleep duration had less than a high school education (36.8% vs. 32.0%) and some college experience (30.0% vs. 27.0%), whereas fewer had a college degree (33.2% vs. 40.9%), with a bias of −16.0% (p < 0.001). Post-matching, educational distributions were well-balanced, with bias reductions across categories (all p-values > 0.01).

Regarding urbanicity, a slightly higher proportion of those with short sleep duration resided in metropolitan areas (71.0%) compared to those with adequate sleep (70.4%), with a bias of 1.4% (p < 0.001). After matching, this difference was further reduced (69.7% vs. 69.4%), with a bias of 0.5% (p = 0.109).

Finally, English language proficiency was slightly higher in those with short sleep duration in the unmatched sample (96.6%) compared to the control group (96.3%), with a bias of 2.0% (p < 0.001). After matching, the English-speaking distribution remained balanced (98.0% vs. 97.9%, bias 0.5%, p = 0.024).

### Association between Sleep Duration and Depression

Table 2 presents the estimated Average Treatment Effects (ATE) of short sleep duration (<7 hours per night) on the incidence of depression. The analysis demonstrates a statistically significant association between insufficient sleep and an increased likelihood of depression.

**Table 2:** Average Treatment Effects of Sleep Duration on Incidence of Depression.

| Outcome | (Treated vs. Control) | Coef. | Std. Err. | z-value | p-value | 95% CI |  |
| --- | --- | --- | --- | --- | --- | --- | --- |
| Depression | Sleep <7hrs. Vs. ≥ 7hrs.) | 0.056 | 0.0048 | 11.79 | 0.000 | 0.047 | 0.065 |
| | Sleep ≥ 7hrs. | 0.194 | 0.0014 | 136.63 | 0.000 | 0.191 | 0.197 |

*This table presents the Average Treatment Effects (ATE) of sleep duration on the incidence of depression. The coefficients (Coef.) represent the estimated average change in depression rate relative to the reference group, where sleep duration $\geq 7$ hrs. Statistical significance is assessed using $z$ -values and $p$ -values, while 95% confidence intervals (CI) indicate the range within which the true effect is expected to lie. Comparisons include respondents with sleep duration < 7 hours and the baseline predicted likelihood of depression among individuals with sleep duration $\geq 7$ hours*

Individuals who reported sleeping less than seven hours per night had a 5.6 percentage-point higher probability of experiencing depression compared to those who met the recommended sleep duration (Coef = 0.056, Std. Err. = 0.0048, z = 11.79, p < 0.001, 95% CI: 0.047 – 0.065).

The high z-value (11.79) and low p-value (<0.001) confirm the robustness of this association, suggesting that short sleep duration significantly increases the risk of depression.

For individuals who reported sleeping at least seven hours per night (reference), the estimated probability of experiencing depression was 19.4% (Coef = 0.194, Std. Err. = 0.0014, z = 136.63, p < 0.001, 95% CI: 0.191 – 0.197). The narrow confidence interval indicates high precision in the estimate, further strengthening the evidence that adequate sleep is associated with a lower likelihood of depression (Table 2).

### Association between Sleep Duration and Mental Health

Table 3 presents the Average Treatment Effects (ATE) of sleep duration on self-reported mental health. The results indicate a significant association between short sleep duration (<7 hours per night) and poorer mental health outcomes.

**Table 3:** Average Treatment Effects of Sleep Duration on Incidence of Mental Health.

| Outcome | (Treated vs. Control) | Coef. | Std. Err. | z-value | p-value | 95% CI |  |
| --- | --- | --- | --- | --- | --- | --- | --- |
| Mental Health | Sleep <7hrs. Vs. ≥ 7hrs.) | 2.24 | 0.1858 | 12.09 | 0.000 | 1.88 | 2.61 |
| | Sleep ≥ 7hrs. | 11.33 | 0.0612 | 185.24 | 0.000 | 11.21 | 11.45 |

*This table presents the Average Treatment Effects (ATE) of sleep duration on the incidence of depression. The coefficients (Coef.) represent the estimated average change in depression rate relative to the reference group, where sleep duration $\geq 7$ hrs. Statistical significance is assessed using $z$ -values and $p$ -values, while 95% confidence intervals (CI) indicate the range within which the true effect is expected to lie. Comparisons include respondents with sleep duration < 7 hours and the baseline predicted likelihood of depression among individuals with sleep duration $\geq 7$ hours.*

Individuals who reported sleeping less than seven hours per night experienced an increase of 2.24 poor mental health days per month compared to those who obtained at least seven hours of sleep (Coef = 2.24, Std. Err. = 0.1858, z = 12.09, p < 0.001, 95% CI: 1.88 – 2.61). The large z-value (12.09) and highly significant p-value (<0.001) confirm the robustness of this association, suggesting that inadequate sleep contributes substantially to worsened mental well-being.

Among individuals who reported sleeping at least seven hours per night, the estimated number of poor mental health days per month was 11.33 days (Coef = 11.33, Std. Err. = 0.0612, z = 185.24, p < 0.001, 95% CI: 11.21 – 11.45). The narrow confidence interval reflects high precision in this estimate, reinforcing the stability of the findings (Table 3).

### Association between Sleep Duration and Physical Health

Table 4 presents the Average Treatment Effects (ATE) of sleep duration on self-reported physical health. The findings indicate a significant association between short sleep duration (<7 hours per night) and poorer physical health outcomes.

**Table 4:**
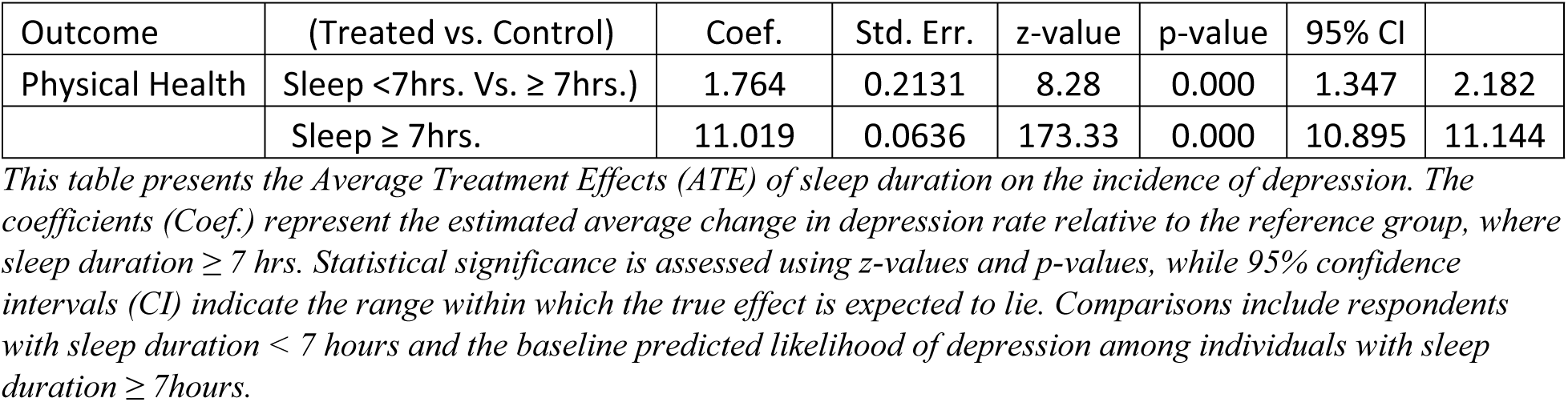
Average Treatment Effects of Sleep Duration on Incidence of Physical Health.

| Outcome | (Treated vs. Control) | Coef. | Std. Err. | z-value | p-value | 95% CI |  |
| --- | --- | --- | --- | --- | --- | --- | --- |
| Physical Health | Sleep <7hrs. Vs. ≥ 7hrs.) | 1.764 | 0.2131 | 8.28 | 0.000 | 1.347 | 2.182 |
| | Sleep ≥ 7hrs. | 11.019 | 0.0636 | 173.33 | 0.000 | 10.895 | 11.144 |

*This table presents the Average Treatment Effects (ATE) of sleep duration on the incidence of depression. The coefficients (Coef.) represent the estimated average change in depression rate relative to the reference group, where sleep duration $\geq 7$ hrs. Statistical significance is assessed using $z$ -values and $p$ -values, while 95% confidence intervals (CI) indicate the range within which the true effect is expected to lie. Comparisons include respondents with sleep duration < 7 hours and the baseline predicted likelihood of depression among individuals with sleep duration $\geq 7$ hours.*

Individuals who reported sleeping less than seven hours per night experienced 1.76 additional poor physical health days per month compared to those who met the recommended sleep duration (Coef = 1.764, Std. Err. = 0.2131, z = 8.28, p < 0.001, 95% CI: 1.347 – 2.182). The large z-value (8.28) and significant p-value (<0.001) confirm the robustness of this association, suggesting that insufficient sleep contributes to poorer physical well-being.

Among individuals who reported sleeping at least seven hours per night, the estimated number of poor physical health days per month was 11.019 days (Coef = 11.019, Std. Err. = 0.0636, z = 173.33, p < 0.001, 95% CI: 10.895 – 11.144) (Table 4).

## DISCUSSION

Our study provides compelling evidence that short sleep duration (<7 hours per night) is significantly associated with adverse mental and physical health outcomes. Specifically, individuals with short sleep experienced a 5.6% increased risk of lifetime depression, in addition to 2.24 more days of poor mental health and 1.8 more days of poor physical health per month compared to those who met the recommended sleep duration. These findings reinforce the growing body of literature highlighting the detrimental effects of insufficient sleep on overall well-being (25, 26).

The relationship between short sleep duration and depression has been well-documented in prior research(31, 32). Consistent with our findings, studies have demonstrated that insufficient sleep is associated with a higher likelihood of developing depression, with sleep disturbances often preceding the onset of depressive symptoms(33, 34). Sleep plays a crucial role in emotional regulation, cognitive processing, and neurobiological stability, and disruptions in sleep patterns can contribute to the development and persistence of mood disorders(35–38). Our results align with previous studies indicating that chronic sleep deprivation alters stress-response mechanisms, particularly through dysregulation of the hypothalamic-pituitary-adrenal (HPA) axis, increasing vulnerability to depression(39–41).

Similarly, the observed association between short sleep duration and increased poor mental health days is consistent with findings suggesting that inadequate sleep exacerbates stress, emotional instability, and decreased resilience to psychological distress(36). Prior research has established that individuals who experience short sleep are more likely to report frequent mental distress, anxiety, and cognitive impairments(25, 42). Our study extends this evidence by quantifying the additional burden of 2.24 more poor mental health days per month, underscoring the substantial impact of insufficient sleep on daily functioning and quality of life.

The connection between short sleep duration and increased poor physical health days is also well-supported by the literature(43). Sleep is integral to physiological restoration, immune function, and metabolic balance, and disruptions in sleep patterns have been linked to higher risks of cardiovascular diseases, obesity, and chronic pain conditions(24, 44–46). Our study found that individuals with insufficient sleep reported an additional 1.8 poor physical health days per month, highlighting the broader implications of sleep deprivation beyond mental health. These results are consistent with evidence showing that sleep deprivation contributes to systemic inflammation, impaired glucose metabolism, and heightened sympathetic nervous system activity, all of which increase susceptibility to chronic physical conditions(47–49).

Several mechanisms may explain the observed relationships between sleep duration and adverse health outcomes. Short sleep duration is known to impair emotional regulation by affecting brain structures such as the prefrontal cortex and amygdala, leading to heightened emotional reactivity and reduced stress tolerance(38, 50, 51). In addition, circadian rhythm disruptions and alterations in neurotransmitter balance (e.g., serotonin and dopamine dysregulation) have been implicated in the development of depressive symptoms among sleep-deprived individuals(52–55).

From a physiological perspective, sleep deprivation has been associated with chronic inflammation, increased oxidative stress, and metabolic dysregulation, which may contribute to both poor mental and physical health outcomes(24, 39, 56). Elevated cortisol levels due to inadequate sleep can further exacerbate mental distress and lead to long-term health consequences, including an increased risk of cardiovascular disease and metabolic disorders(57, 58).

### Public Health Implications

Our findings have significant public health and policy implications, emphasizing the need for interventions aimed at promoting healthy sleep habits. Given the high prevalence of short sleep duration in the United States, particularly among working adults, healthcare providers should integrate sleep assessments into routine medical evaluations and advocate for strategies to improve sleep hygiene. Workplace policies that encourage flexible work schedules, reduced shift work burden, and stress management programs may help mitigate the adverse effects of insufficient sleep on mental and physical health.

Furthermore, public health campaigns should emphasize the importance of achieving at least seven hours of sleep per night, particularly for high-risk populations such as shift workers, individuals with high stress levels, and those with pre-existing health conditions. Sleep interventions, including cognitive-behavioral therapy for insomnia (CBT-I), relaxation techniques, and digital tools for sleep tracking, could serve as effective strategies for improving sleep duration and quality.

### Strengths and Limitations

This study has several strengths that enhance its validity and generalizability. By utilizing a large, nationally representative dataset, our findings are applicable to a broad population. The use of Inverse Probability Weighting (IPW) helped minimize confounding biases and allowed for a robust estimation of the causal effects of short sleep duration on depression, mental health, and physical health. Additionally, our study accounted for sociodemographic factors, employment status, and urbanicity, further strengthening the reliability of our results.

However, several limitations must be acknowledged. First, our study relied on self-reported sleep duration, which may be subject to recall bias and reporting inaccuracies. Objective sleep measures, such as actigraphy or polysomnography, would provide more precise estimates of sleep patterns. Second, the cross-sectional nature of our study prevents causal inference, as bidirectional relationships may exist between sleep duration and health outcomes. For example, while short sleep duration may contribute to depression, depressive symptoms may also lead to sleep disturbances, creating a cyclical relationship that cannot be fully disentangled in this analysis. Finally, unmeasured confounders, such as genetic predispositions, chronic stress levels, and dietary factors, may have influenced the observed associations. Future research utilizing longitudinal designs and intervention-based studies would be beneficial in further elucidating the causal pathways linking sleep duration to mental and physical health outcomes.

### Conclusion

In conclusion, this study demonstrates that short sleep duration (<7 hours per night) is significantly associated with a higher incidence of depression, more poor mental health days, and increased poor physical health days. These findings highlight the critical role of adequate sleep in maintaining both mental and physical well-being and underscore the need for public health interventions aimed at promoting healthy sleep habits. Addressing sleep deprivation at the population level through educational campaigns, workplace modifications, and clinical interventions could help mitigate the adverse effects of insufficient sleep and improve overall health outcomes.

## Data Availability

All relevant data are within the manuscript and its supporting information files.

